# Altered White Matter Tracts in Bipolar Disorder: Insights from DTI Analysis

**DOI:** 10.1101/2025.06.03.25328854

**Authors:** Ayda Mostafavi, Hanieh Mohammadizadeh, Golnaz Younesi, Ebrahim Rahmani

## Abstract

**Introduction:** Bipolar Disorder (BD) is characterized by marked disruptions in emotional regulation and cognitive control, often accompanied by structural abnormalities in the brain’s white matter. Diffusion tensor imaging (DTI) offers a window into white matter microstructure through metrics such as fractional anisotropy (FA), mean diffusivity (MD), radial diffusivity (RD), and axial diffusivity (AD). This study aimed to examine white matter tract alterations in individuals with BD compared to healthy controls (HCs) and to explore potential associations between DTI metrics and clinical symptom severity.

**Methods:** We conducted a voxel-wise whole-brain DTI analysis in a sample of BD patients and age-matched HCs, focusing on FA, MD, RD, and AD values. Group comparisons were performed to identify significant differences in white matter integrity. In addition, we assessed correlations between DTI metrics and psychological assessment scores to investigate links between structural alterations and clinical features.

**Results:** Compared to HCs, BD patients exhibited significantly lower FA in several major white matter tracts, including the cingulum (olfactory tract), forceps minor of the corpus callosum, fornix, inferior fronto-occipital fasciculus, and superior longitudinal fasciculus. These reductions suggest disrupted microstructural coherence. In parallel, elevated MD, RD, and AD in overlapping regions point to possible myelin degeneration or axonal injury. However, associations between DTI metrics and psychological symptom scores were weak and did not reach statistical significance.

**Discussion/Conclusion:** These findings reinforce the presence of widespread white matter abnormalities in BD, particularly in tracts relevant to emotional and cognitive processing. While structural disruptions were evident, their weak correlations with symptom severity highlight the complex relationship between brain microstructure and clinical expression in BD. Future studies are warranted to clarify the diagnostic and prognostic relevance of DTI-based biomarkers in mood disorders.

## Introduction

Bipolar disorder (BD) is a chronic psychiatric illness marked by episodes of mania, hypomania, and depression (Jonathan et al., 2021; Patel et al., 2018). It affects about 1–2% of the global population (Merikangas et al., 2011; Vieta et al., 2018) and has a heritability estimate of 60– 80% (McGuffin et al., 2003). Although white matter (WM) abnormalities have been reported in BD, findings remain inconsistent across imaging modalities and populations (Benedetti et al., 2011; Magioncalda et al., 2016). Most studies have not used multiple DTI-derived measures or included BD patients with psychiatric comorbidities.

This study addresses that gap by examining WM tract alterations in BD using several DTI indices (e.g., FA, MD, RD, QA, NQA) and including comorbid presentations. This **multi-index and comorbidity-inclusive design** enhances understanding of WM changes in BD and may clarify neural markers relevant for early detection and intervention.

White matter (WM) refers to myelinated axonal fibers that facilitate communication between different brain regions, enabling coordinated neural activity and functional integration (Safaiyan et al., 2021). Structural integrity of WM is essential for cognitive and emotional processing, both of which are often impaired in bipolar disorder (BD). BD is classified into two major subtypes: Type I, which involves at least one full manic episode, and Type II, characterised by hypomania and major depressive episodes (Vieta et al., 2018). Recent findings suggest that alterations in WM tracts may contribute to the pathophysiology of both subtypes (Strakowski et al., 2012). Impaired WM microstructure may underlie the cognitive deficits and affective instability seen in BD patients, particularly during active phases of the illness (Liu et al., 2017).

The prevalence of psychiatric comorbidities in BD has been subjected to various types of research in the past. Yet, investigation of alteration in the WM tracts as a possible cause of BD and psychiatric comorbidities such as depression and ADHD are almost unique to this study. To measure WM tracts different Neuroimaging techniques are used. Neuroimaging provides the opportunity to explore the pathophysiology of disorders and their neural mechanisms (van Ewijk et al., 2012). However, diffusion tensor imaging (DTI) has been proven to be a very reliable representation of such techniques. Based on prior studies, not only WM integrity could be altered in patients with BD (Benedetti et al.,2011), but also a correlation between white matter alterations and bipolar disorder has been observed (Marlinge et al., 2014). Although reduction of regional brain volume in bipolar type I is shown in various researches, WM alteration is accrued more in bipolar type II (Maller et al., 2014). Furthermore, alternations in WM among BD type I indicated distinct spatial patterns in different stages of the disease. These patterns mostly affect the active phases and correlate to some cognitive disabilities. It is proposed that there is a complex characteristic and state-dependent pathogenesis of abnormality of WM in BD patients (Magioncalda et al.,2016).

The observations made reflect upon the fact that the WM and corpus callosum (CC) volumes were significantly smaller in BD patients in the early and late stages compared to the healthy group (Duarte et al., 2018). Investigations show that there is a link between cognitive function and flexibility to the WM abnormalities in bipolar disorder patients (Oertel-Knöchel et al., 2014; Radoeva et al., 2020).

Literature suggests that the abnormalities in regional brain thickness and white matter integrity in BD patients, and some differences between bipolar type I and type II have been observed. It was found that WM microstructural abnormality is a common feature in both psychotic and non-psychotic bipolar disorder (Lee et al., 2022). BD onset can be traced back to the WM disruptions (Linke et al., 2020) and information flow via different brain regions through WM tracts. Hence, WM tract alteration might cause altered emotional and cognitive processing in patients (Stupar, 2010; Tanrıkulu et al., 2022). Although structural abnormalities in bipolar patients have been demonstrated by several neuroimaging examinations (Ellison-Wright & Bullmore, 2010), they are not indicative of detailed neural mechanisms of BD. A thorough understanding of symptoms severity biomarkers can thus enable early risk detection and better interventions for bipolar patients (Santos et al., 2022).

Studying changes in white matter (WM) microstructure through diffusion tensor imaging (DTI) has offered important insights into the neural underpinnings of BD. However, previous findings on fractional anisotropy (FA) alterations remain inconsistent, with reports of reduced (e.g., Benedetti et al., 2011; Barnea-Goraly et al., 2009), increased (e.g., Haznedar et al., 2005), or no significant differences (e.g., Beyer et al., 2005) in BD patients compared to healthy controls. Recent large-scale studies (e.g., Favre et al., 2021; Cao et al., 2024) continue to reveal variability in WM abnormalities depending on clinical characteristics, symptom profiles, and comorbidities, rather than strictly BD subtype.

What makes the current study unique is its use of multiple DTI-derived indices—FA, MD, RD, QA, and NQA—to evaluate WM tract alterations, as well as its inclusion of BD patients with psychiatric comorbidities, such as ADHD and depression. By moving beyond a simple BD type I vs. type II classification and incorporating symptom-based differentiation, this study aims to provide a more nuanced, transdiagnostic understanding of white matter pathology in BD. This multi-index, symptom-informed approach may clarify inconsistent findings in the literature and offer novel biomarkers for future clinical application.

## Materials and Methods

### Participants

The study sample comprised 49 patients with BD and 130 HCs which was obtained from the UCLA Consortium for Neuropsychiatric Phenomics (CNP) dataset through OpenNeuro with the access number ds000030 (1,2). The CNP contains neuroimaging data, neuropsychological assessments, and neurocognitive tasks for both healthy people and those with neuropsychiatric disorders (1). Subjects were adults aged 21-50 who studied 8 years or more at school, had no history of any other medical issue and had a negative urine test for substance abuse <u>(2).</u> Participants who were left-handed, pregnant and experienced head trauma with loss of consciousness and other contraindications to MRI scanning were excluded from the imaging study (1). Exclusion criteria for the healthy group were: a diagnosis of Psychiatric Disorder, Substance Abuse/Dependence, and Bipolar Disorder, a current diagnosis of major depressive disorder, suicide attempt, anxiety disorder, and Deficit Hyperactivity Disorder (ADHD) (2). Participants provided written informed consent based on UCLA’s Institutional Review Board after receiving a verbal explanation about the study (1).

### BD Diagnosis and Assessment

All psychiatric diagnoses in this study were made according to the Diagnostic and Statistical Manual of Mental Disorders, Fourth Edition (DSM-IV), which was the standard at the time of data collection (2007–2011) (Poldrack et al., 2016). The diagnosis of bipolar disorder (BD) was confirmed by licensed clinical psychiatrists using the Structured Clinical Interview for DSM-IV (SCID-I). Additional comorbid conditions such as schizophrenia and ADHD were also diagnosed using DSM-IV criteria and standardized clinical interviews. The Adult ADHD Interview was used to assess attention-deficit/hyperactivity disorder, and the Hamilton Depression Rating Scale (HAM-D) was administered to measure depressive symptom severity in participants with mood-related disorders (Hamilton, 1960). This structured, multi-scale diagnostic approach ensures clinical validity and consistency within the dataset.

### Pre-processing

Data were pre-proceed using the Statistical Parametric Mapping (SPM) software package (SPM v.12, “http://www.fill.ion.ucl.ac.uk/spm/software/spam12”) and its Computational Anatomy Toolbox (CAT12, “http://www.neuro.uni-jena.de/cat/”) based on CAT12 manual (“http://www.neuro.uni-jena.de/cat12/CAT12-Manual.pdf”). First, we used the DARTEL registration template (1) for spital registration of T1 images to the Montreal Neurological Institute (MNI). Individuals’ brain structure data were divided into cerebrospinal fluid (CSF), gray matter (GM), and WM. Then we used the projection-based thickness method for the surface reconstruction of every person (2). Reconstruction of the central cortical surface involved topology correction, spherical inflation, and registration. Then we utilized the central surface to calculate the gyrification index (GI), the fractal dimension (FD), and the values of sulcal depth (SD). At last, the surface measurements of the merged left and right hemispheres were resampled onto the 32k mesh resolution. Data quality was inspected in two steps: 1) before pre-processing, images were checked out for artifacts by visual inspection. 2) After segmentation, image quality and sample homogeneity of all images were checked by using inhomogeneity contrast, noise contrast, and root mean square of voxel resolution natively implemented in the CAT12 (“http://dbm.neuro.uni-jena.de/HBM2013/Dahnke02.pdf”). No one was excluded for artifacts.

Then spherical harmonic reconstructions method (3) was used to analyze the estimation and comparison of cortical surface complexity (2). Briefly, we extracted the spherical harmonic coefficients up to a maximum l-value (or degree) with ranges of 11-29 for every vortex. Then we used a modified fast Fourier transform technique to remap it into the harmonic space. We also calculated cortical complexity as a slope by regressing log (area) versus the max l-value. Then, using spherical mesh in the FreeSurfer software suite, we re-parametrized the local cortical complexity in a common coordinate system (“https://surfer.nmr.mgh.harvard.edu/”). After surface data were resampled for Fractal Dimension, we smoothed them using 25 mm (and 20 mm) full width at half maximum (FWHM) Gaussian kernel prior to 2nd-level analysis. Finally, to reach Statistical significance, family-wise error (FWE) correction was performed, and a threshold of P < 0.05 was considered significant.

### Data Acquisition

The data was extracted from www.open-datacommons.org, number ds000030, revision 1.0.3 (9). Standard processing (FMRIPREP version 0.4.4) and statistical modeling (ANTs N4BiasFieldCorrection v2.1.0) have been implemented to make the CNP database easier to use for scientific proposes and increase its open availability. Denoising was not conducted to leave more flexibility for the following research, although potential confound regressors were obtained for every trial (10).

The registered participant underwent several behavioural tests, including Demographics & General Health, Symptoms, Traits, Neurocognitive Tasks, and Neuropsychological Assessment (11). The total number of subjects was 272, including 130 healthy and 142 diagnosed with disorders.

### Neuroimaging preprocessing

Diffusion tensor imaging (DTI) data preprocessing was conducted using FSL (FMRIB Software Library; Smith et al., 2004), following standard pipelines to ensure artifact removal and robust diffusion metric estimation. First, all diffusion-weighted volumes were corrected for eddy current distortions and head motion using the eddy_correct tool, and the non-brain tissue was removed with the Brain Extraction Tool (BET). Next, diffusion tensors were fitted at each voxel using linear least squares estimation, generating fractional anisotropy (FA), mean diffusivity (MD), radial diffusivity (RD), and axial diffusivity (AD) maps. These individual diffusion maps were non-linearly registered to a standard FA template in MNI152 space using Tract-Based Spatial Statistics (TBSS). The FA images were aligned to a common target, averaged to create a mean FA skeleton, and each subject’s FA data was projected onto this skeleton for voxel-wise analysis. A similar pipeline was followed for the remaining DTI metrics (MD, RD, AD). Finally, quality assurance (QA) procedures were implemented to detect and exclude datasets with significant motion artifacts, signal dropout, or misalignment.

### Statistical Analysis

We performed chi-square tests and independent samples t-tests for parametric tractographic and demographic data, and the Mann–Whitney U test for non-parametric data, using IBM SPSS Statistics (version XX). Tractographic indices of white matter were compared among the control, bipolar, and ADHD groups using independent t-tests for normally distributed data and the Mann–Whitney U test for non-normal distributions. To correct for multiple comparisons, we used the Benjamini–Hochberg method. A p-value of less than 0.05 was considered statistically significant..

### Findings

**Table 1** presents data on demographic characteristics and neuropsychiatric assessment scores comparing individuals with bipolar disorder (BPD) (n=48) and a healthy control group (n=114). In terms of neuropsychiatric assessment scores, the BPD group exhibited moderate levels of manic symptoms (mean YMRS score: 11.91) and depressive symptoms (mean HAMD 28 score: 18.58). They also reported a higher presence of ADHD symptoms (mean ASRS score: 13.25) compared to the healthy control group (mean ASRS score: 8.64). Furthermore, the BPD group had higher scores in all subcategories of the BPRS, indicating greater severity of mania, negative symptoms, positive symptoms, and depression/anxiety symptoms. Similarly, they had higher scores in all subcategories of the HSCL, reflecting increased levels of somatization, obsessive-compulsive symptoms, interpersonal sensitivity, depression, and anxiety.

**Table 1.** Demographics and neuropsychiatric assessment scores in BPD patients and healthy controls.

| Demographic characteristics | HC (n=114) | BPD (n=48) | p-value |
| --- | --- | --- | --- |
| Age (Years) |  |  |  |
| Gender (Male) |  |  |  |
| Education (Years) |  |  |  |
| YMRS | NA | 11.91 (11.1) | 0 |
| HAMD 28 | NA | 18.58 (13.2) | 0 |
| ASRS | 8.64 (2.91) | 13.25 (4.92) | 3.679 |
| **BPRS** |  |  |  |
| Mania | NA | 12.14 (4.81) | 0 |
| Negative symptom | NA | 5.62 (2.14) | 0 |
| Positive symptom | NA | 6.83 (2.11) | 0 |
| Depression/anxiety | NA | 11.68 (4.97) | 0 |
| Total | NA | 44.69 (11.14) | 0 |
| **HSCL** |  |  |  |
| Somatization | 2.48 (2.7) | 7.75 (7.3) | 3.061 |
| Obsessive-compulsive | 3.94 (3.4) | 9.27 (5.82) | 8.232 |
| Interpersonal sensitivity | 2.56 (2.29) | 7.66 (5.07) | 1.209 |
| Depression | 3.86 (3.8) | 10.5 (6.95) | 5.151 |
| Anxiety | 1.26 (1.69) | 4.47 (3.76) | 5.383 |
| Global severity | 18.42 (13.71) | 50.5 (31.99) | 5.223 |

\*ANCOVA controlled for age, gender, and education.
BPD: Bipolar disorder, ASRS: Adult ADHD Self-Report Scale, BPRS: Brief Psychiatric Rating Scale, HAMD 28: Hamilton depression rating scale, HC: healthy control, HSCL: Hopkins Symptom Checklist, YMRS: Young Mania Rating Scale

Based on the provided information and findings in table 2, there appears to be a link between white matter tracts and bipolar disorder. Significant differences were observed in various white matter tracts between the HC group and the BPD group, as indicated by differences in fractional anisotropy (FA), mean diffusivity (MD), radial diffusivity (RD), and axial diffusivity (AD). The BPD group exhibited lower FA in several tracts, including the cingulum of the olfactory tract, corpus callosum (body, forceps major, and forceps minor), fornix, inferior fronto-occipital fasciculus, and superior longitudinal fasciculus 1. These findings suggest disrupted microstructural integrity and compromised white matter connectivity in these specific regions associated with bipolar disorder. Additionally, higher MD, RD, and AD were observed in the fornix, cingulum of the olfactory tract, corpus callosum (forceps minor), and inferior fronto-occipital fasciculus in the BPD group, indicating potential abnormalities in myelin or axonal properties. These findings collectively support the notion that alterations in white matter tracts are associated with BPD, potentially contributing to cognitive, emotional, and functional impairments commonly observed in this psychiatric condition.

**Table 2.**
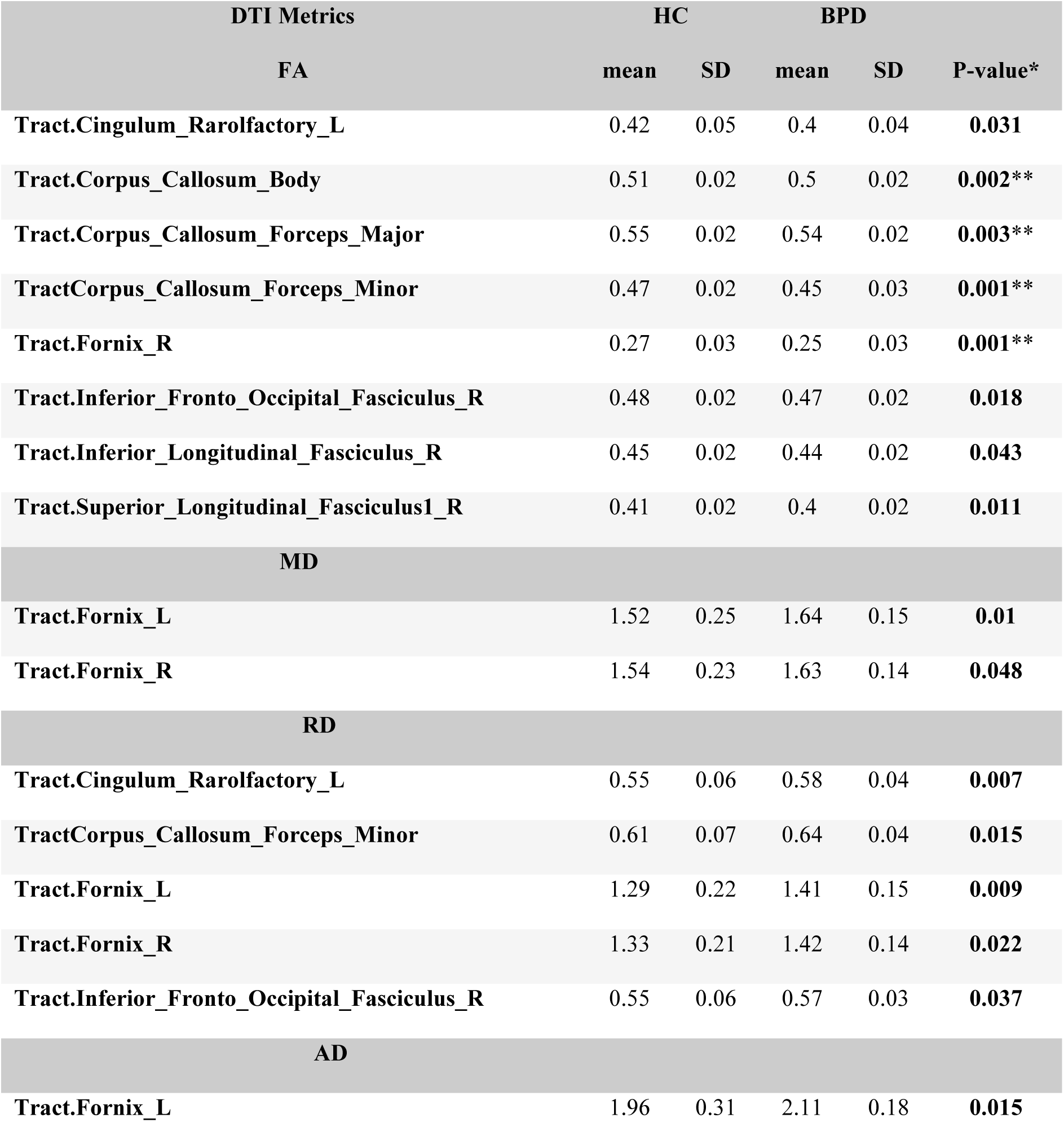
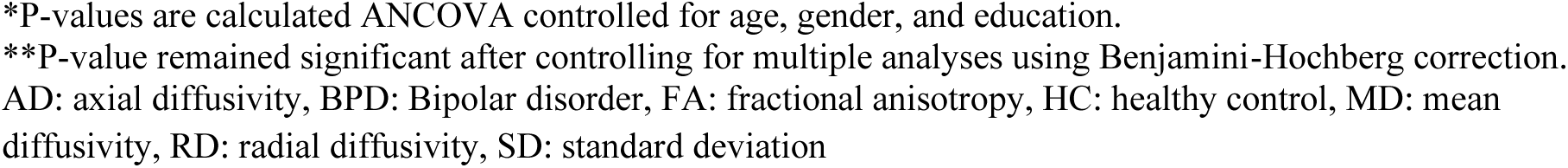
WM tract with significant differences between HC and BPD groups.

| DTI Metrics | HC |  | BPD |  | P-value* |
| --- | --- | --- | --- | --- | --- |
|  | mean | SD | mean | SD |  |
| **FA** |  |  |  |  |  |
| Tract.Cingulum_Rarolfactory_L | 0.42 | 0.05 | 0.4 | 0.04 | **0.031** |
| Tract.Corpus_Callosum_Body | 0.51 | 0.02 | 0.5 | 0.02 | **0.002\*\*** |
| Tract.Corpus_Callosum_Forceps_Major | 0.55 | 0.02 | 0.54 | 0.02 | **0.003\*\*** |
| TractCorpus_Callosum_Forceps_Minor | 0.47 | 0.02 | 0.45 | 0.03 | **0.001\*\*** |
| Tract.Fornix_R | 0.27 | 0.03 | 0.25 | 0.03 | **0.001\*\*** |
| Tract.Inferior_Fronto_Occipital_Fasciculus_R | 0.48 | 0.02 | 0.47 | 0.02 | **0.018** |
| Tract.Inferior_Longitudinal_Fasciculus_R | 0.45 | 0.02 | 0.44 | 0.02 | **0.043** |
| Tract.Superior_Longitudinal_Fasciculus1_R | 0.41 | 0.02 | 0.4 | 0.02 | **0.011** |
| **MD** |  |  |  |  |  |
| Tract.Fornix_L | 1.52 | 0.25 | 1.64 | 0.15 | **0.01** |
| Tract.Fornix_R | 1.54 | 0.23 | 1.63 | 0.14 | **0.048** |
| **RD** |  |  |  |  |  |
| Tract.Cingulum_Rarolfactory_L | 0.55 | 0.06 | 0.58 | 0.04 | **0.007** |
| TractCorpus_Callosum_Forceps_Minor | 0.61 | 0.07 | 0.64 | 0.04 | **0.015** |
| Tract.Fornix_L | 1.29 | 0.22 | 1.41 | 0.15 | **0.009** |
| Tract.Fornix_R | 1.33 | 0.21 | 1.42 | 0.14 | **0.022** |
| Tract.Inferior_Fronto_Occipital_Fasciculus_R | 0.55 | 0.06 | 0.57 | 0.03 | **0.037** |
| **AD** |  |  |  |  |  |
| Tract.Fornix_L | 1.96 | 0.31 | 2.11 | 0.18 | **0.015** |

\*P-values are calculated ANCOVA controlled for age, gender, and education.
\*\*P-value remained significant after controlling for multiple analyses using Benjamini-Hochberg correction.
AD: axial diffusivity, BPD: Bipolar disorder, FA: fractional anisotropy, HC: healthy control, MD: mean diffusivity, RD: radial diffusivity, SD: standard deviation

The key findings in **table 3** show weak correlations between these DTI metrics and psychological scores. Specifically, FA and MD exhibit weak positive correlations with the scales, suggesting that better white matter integrity or increased diffusion might be associated with higher symptom severity. In contrast, RD and AD show weak negative correlations, indicating that increased diffusion might be linked to milder symptoms. However, all these correlations are not statistically significant, meaning they may not be meaningful or reliable. Overall, the lack of significant findings suggests that any observed relationships between white matter integrity and psychological symptoms should be interpreted with caution, as they may not reflect true associations.

**Table 3:** correlations between these DTI metrics and psychological scores.

| DTI metrics | Statistic | ASRS | HAMD 28 | YMRS |
| --- | --- | --- | --- | --- |
| **FA** |  |  |  |  |
| **Tract.Cingulum_Rarolfactory_L** | B | 0.068 | 0.173 | 0.155 |
|  | P | 0.640 | 0.250 | 0.295 |
|  | PI | 0.954 | NA | NA |
| **Tract.Corpus_Callosum_Body** | B | -0.038 | 0.012 | 0.089 |
|  | P | 0.790 | 0.936 | 0.543 |
|  | PI | 0.680 | NA | NA |
| **Tract.Corpus_Callosum_Forceps_Major** | B | 0.023 | 0.076 | 0.090 |
|  | P | 0.873 | 0.611 | 0.540 |
|  | PI | 0.750 | NA | NA |
| **TractCorpus_Callosum_Forceps_Minor** | B | 0.011 | -0.02 | -0.139 |
|  | P | 0.933 | 0.890 | 0.323 |
|  | PI | 0.863 | NA | NA |
| **Tract.Fornix_R** | B | -0.104 | -0.139 | 0.031 |
|  | P | 0.543 | 0.445 | 0.869 |
|  | PI | 0.923 | NA | NA |
| **Tract.Inferior_Fronto_Occipital_Fasciculus_R** | B | -0.016 | -0.091 | -0.048 |
|  | P | 0.904 | 0.532 | 0.738 |
|  | PI | 0.165 | NA | NA |
| **Tract.Inferior_Longitudinal_Fasciculus_R** | B | -0.081 | -0.029 | -0.032 |
|  | P | 0.578 | 0.845 | 0.825 |
|  | PI | 0.398 | NA | NA |
| **Tract.Superior_Longitudinal_Fasciculus1_R** | B | 0.043 | 0.034 | 0.110 |
|  | P | 0.769 | 0.825 | 0.474 |
|  | PI | 0.382 | NA | NA |
| **MD** |  |  |  |  |
| **Tract.Fornix_L** | B | -0.184 | 0.156 | 0.020 |
|  | P | 0.213 | 0.290 | 0.895 |
|  | PI | 0.840 | NA | NA |
| **Tract.Fornix_R** | B | 0.068 | -0.110 | -0.175 |
|  | P | 0.686 | 0.538 | 0.349 |
|  | PI | 0.879 | NA | NA |
| **RD** |  |  |  |  |
| **Tract.Cingulum_Rarolfactory_L** | B | -0.178 | -0.270 | -0.219 |
|  | P | 0.193 | 0.054 | 0.113 |
|  | PI | 0.998 | NA | NA |
| **TractCorpus_Callosum_Forceps_Minor** | B | 0.024 | 0.089 | 0.132 |
|  | P | 0.856 | 0.527 | 0.334 |
|  | PI | 0.542 | NA | NA |
| **Tract.Fornix_L** | B | -0.180 | 0.155 | 0.004 |
|  | P | 0.239 | 0.309 | 0.979 |
|  | PI | 0.823 | NA | NA |
| **Tract.Fornix_R** | B | 0.092 | -0.064 | -0.156 |
|  | P | 0.582 | 0.716 | 0.403 |
|  | PI | 0.878 | NA | NA |
| **Tract.Inferior_Fronto_Occipital_Fasciculus_R** | B | 0.033 | 0.090 | 0.046 |
|  | P | 0.804 | 0.528 | 0.744 |
|  | PI | 0.375 | NA | NA |
| **AD** |  |  |  |  |
| **Tract.Fornix_L** | B | -0.175 | 0.144 | 0.044 |
|  | P | 0.246 | 0.341 | 0.774 |
|  | PI | 0.873 | NA | NA |

AD: axial diffusivity, ASRS: adult ADHD self-report scale, DTI: diffusion tensor imaging, FA: fractional anisotropy, HAMD 28: Hamilton depression rating scale, MD: mean diffusivity, PI: P-value of interaction, RD: radial diffusivity, YMRS: young mania rating scale

The study, as shown in **table 4**, found weak associations between DTI metrics (FA, MD, RD, AD) and various symptoms, but none of these associations reached statistical significance. This implies that the DTI metrics examined may not be reliable predictors of the symptoms investigated in this particular study.

**Table 4:**
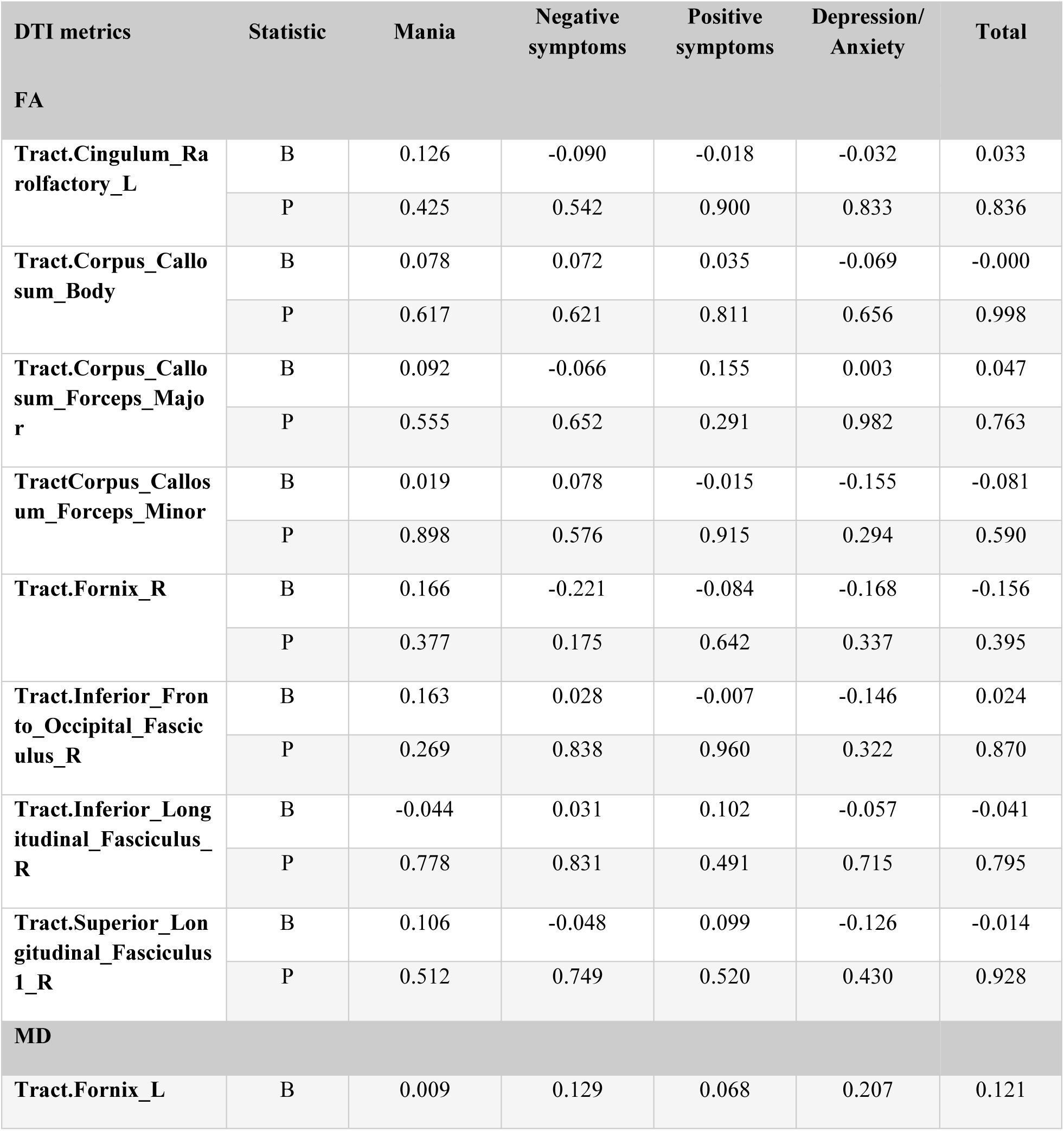
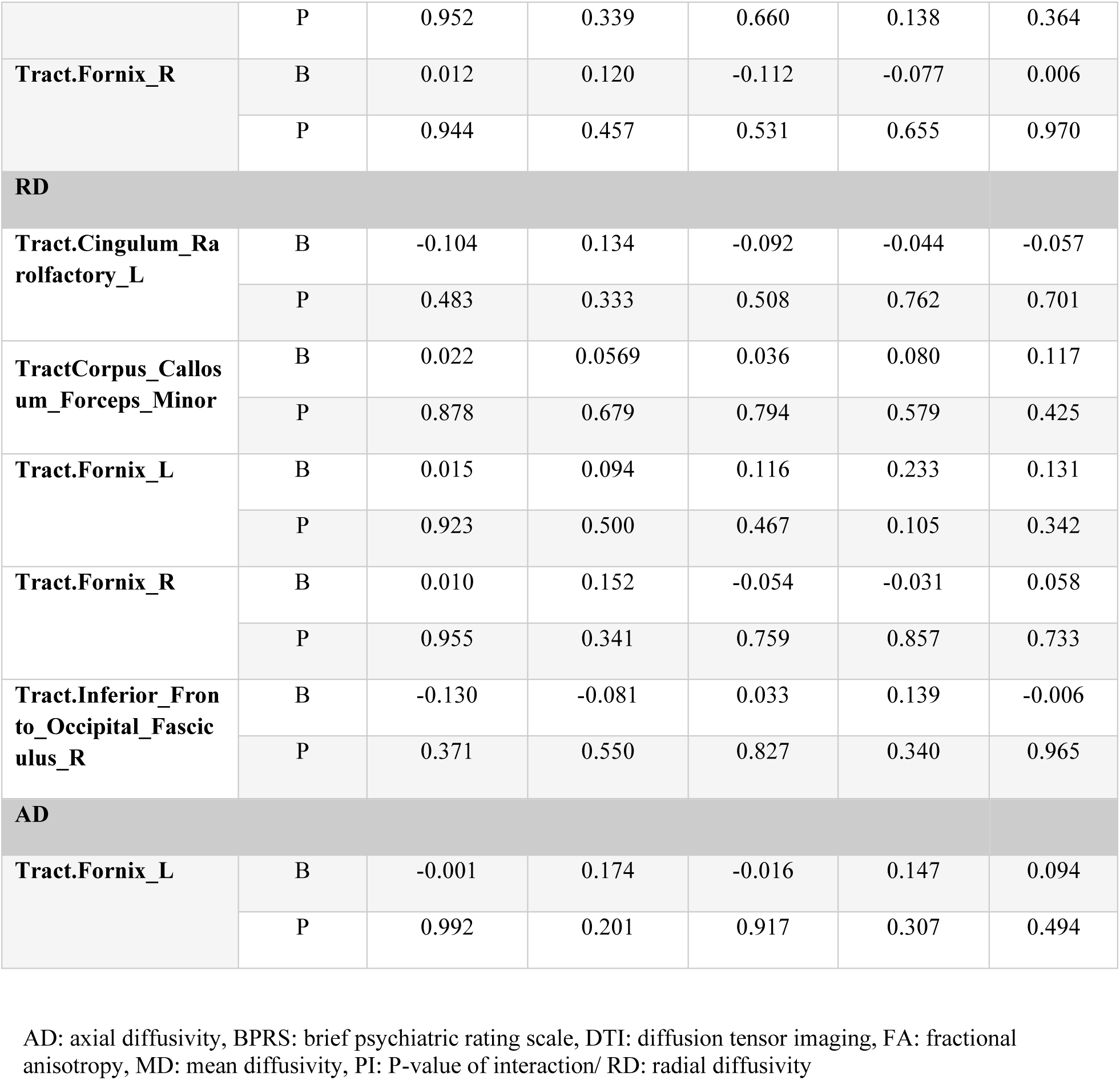
Association between significant tracts and BPRS scores.

Across different tracts analyzed, FA, MD, RD, and AD show negative associations with psychological symptoms, indicating that higher FA and lower MD, RD, and AD values are generally linked with lower symptom severity (**table 5**). However, many of these associations do not reach statistical significance, suggesting that they may not be robust or consistent across different tracts and symptom subdomains. Further research with larger sample sizes and more sophisticated statistical analyses may help elucidate the precise nature of these associations and their potential clinical implications for understanding and treating psychological disorders.

**Table 5:**
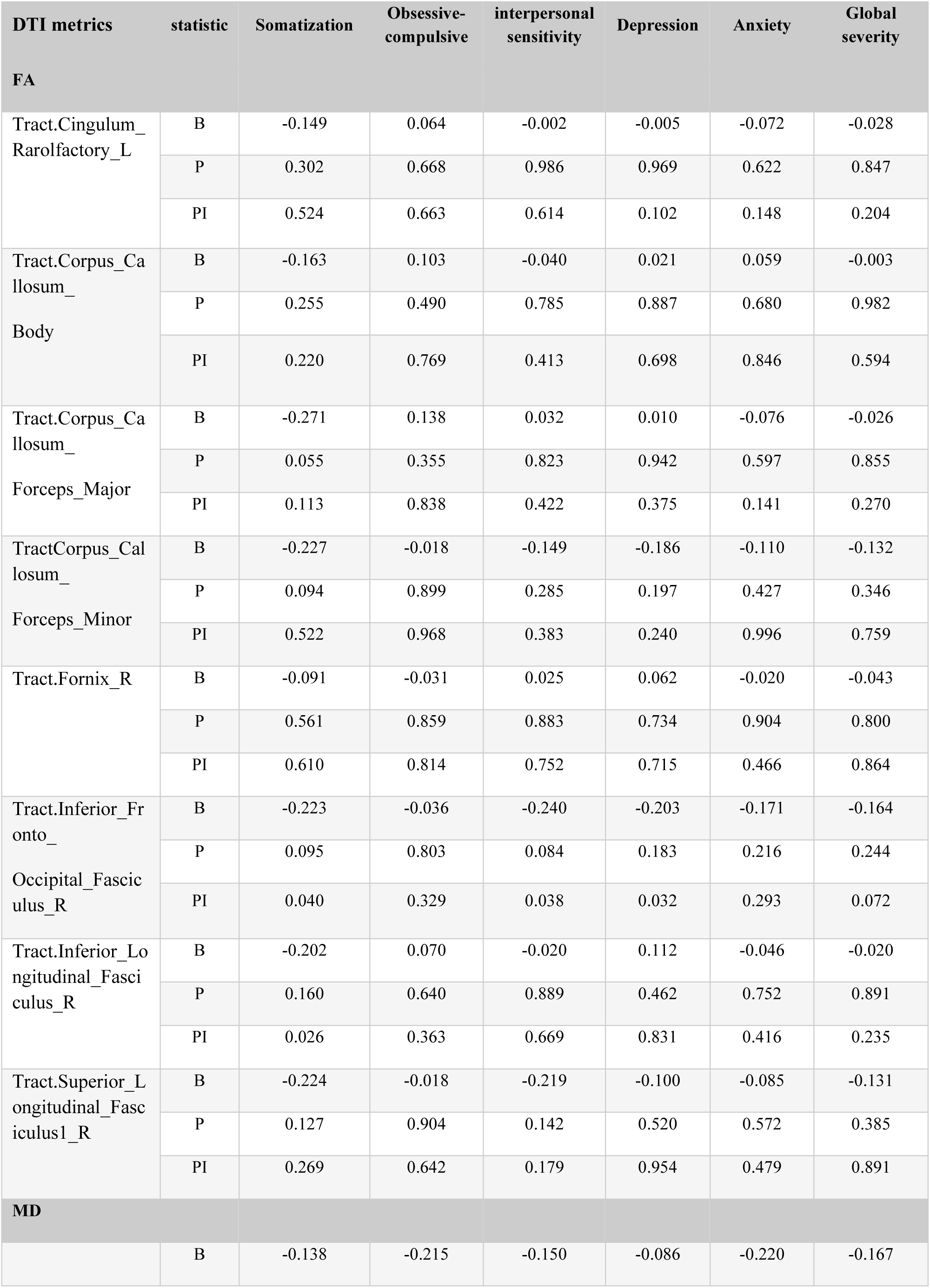
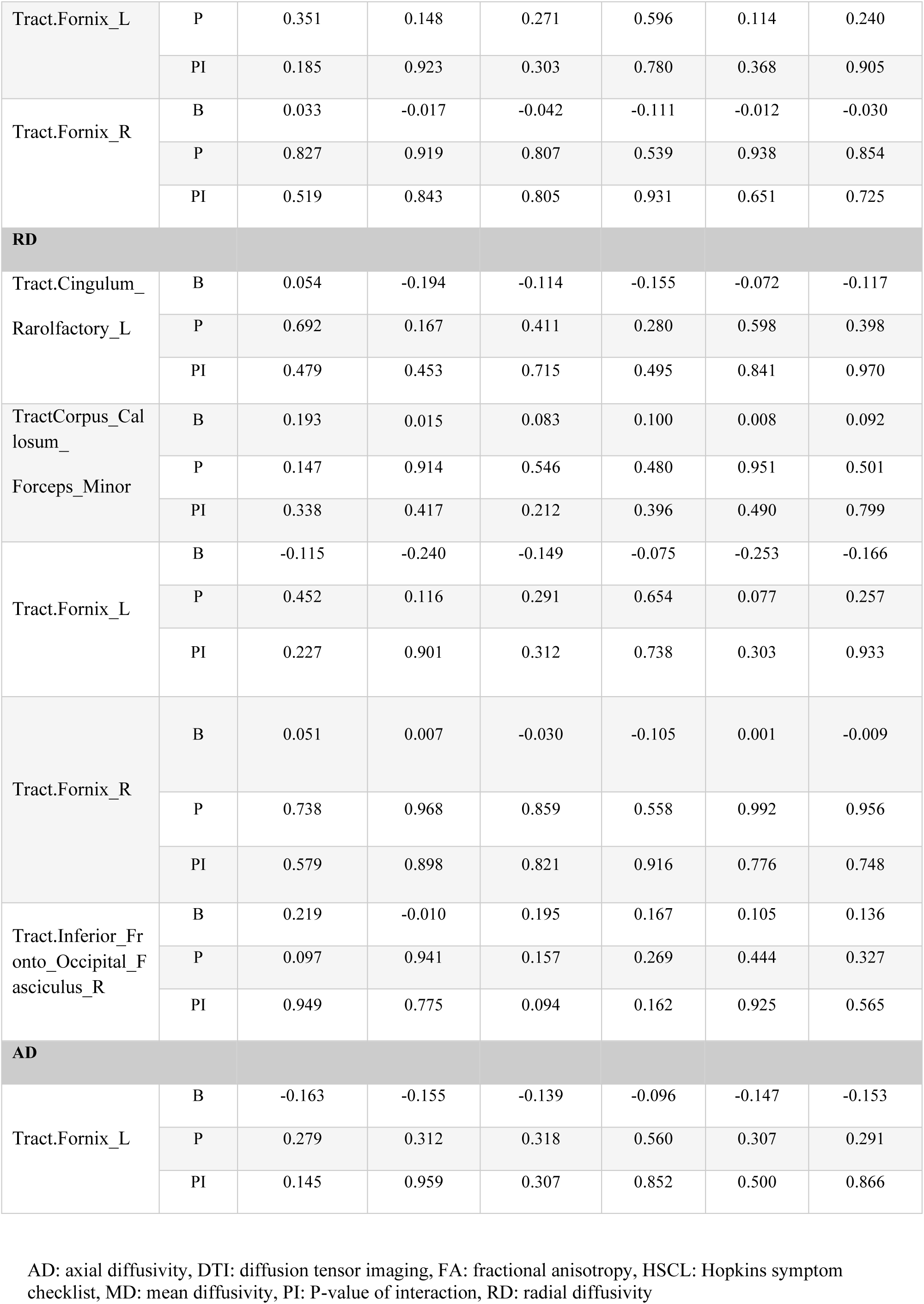
tracts & associations with psychological symptoms.

## Discussion

Using the UCLA CNP dataset, authors sought to provides a connectometry study investigating the associations between white matter tract alterations in patients suffering from BPD compared to HCs.

### Demographic and Neuropsychiatric Assessment Scores

The demographic and neuropsychiatric assessment in our study yielded significant differences between BPD and HC groups. The BPD group demonstrated moderate manic symptoms with a mean Young Mania Rating Scale (YMRS) score of 11.91 and notable depressive symptoms with a mean Hamilton Depression Rating Scale (HAMD 28) score of 18.58.

In a cross-sectional study by Li et al. (1) examining the YMRS and HAMD 28 scores among BPD, major depressive disorder (MDD), and healthy controls (HC), distinct patterns were observed across these groups. YMRS scores were significantly higher in both depressed and euthymic bipolar patients (d-BD: 2.59 ± 2.75, e-BD: 1.73 ± 2.30) compared to euthymic major depressive disorder patients (e-MDD: 0.22 ± 0.61) and HCs (HC: 1.01 ± 1.95), indicating greater manic symptoms in the bipolar groups (F = 7.48, P < 0.001). Similarly, HAMD scores were markedly elevated in depressive bipolar and major depressive disorder patients (d-BD: 18.21 ± 6.42, d-MDD: 17.04 ± 5.96) compared to euthymic bipolar (e-BD: 2.07 ± 2.10), euthymic major depressive disorder (e-MDD: 2.71 ± 2.19), and HCs (HC: 0.39 ± 1.10), reflecting higher depression levels in the depressive states (F = 283.61, P < 0.001). These findings highlight the greater severity of both manic and depressive symptoms in BPD compared to major depressive disorder and HCs.

Additionally, BPD patients were found to have a higher prevalence of ADHD symptoms in our study, as evidenced by an elevated Adult ADHD Self-Report Scale (ASRS) score of 13.25 compared to 8.64 in HCs.

In a study by Ertek et al (2) involving 1517 cases, the YMRS and HAMD 28 scores were compared between BPD and HCs. Their findings revealed that 8.6% of the participants had a bipolar severity score of 40 or higher, while 48% had a bipolar frequency score of 18 or above. Additionally, the Adult ADHD Self-Report Scale (ASRS) demonstrated that 3.7% of cases scored 49 or higher, indicating a significant presence of ADHD.

When exploring the associations between ADHD and BPD, 70.4% of participants with ADHD reflected a high bipolar frequency score, compared to 47.2% of those without ADHD. Additionally, 37.0% of participants with ADHD demonstrated high bipolar severity scores, versus 7.6% in lack thereof. These differences were found to be statistically significant, underscoring a strong association between ADHD and increased bipolar symptoms. Furthermore, patients with ADHD had significantly higher rates of psychiatric diseases (23.6% vs. 7.6%), forensic events (14.5% vs. 1.6%), and traffic accidents (18.2% vs. 9.0%) compared to lack thereof (2). These findings highlight the complex interplay between ADHD and bipolar symptoms, with ADHD potentially intensifying the severity and frequency of bipolar manifestations. Our findings highlight the complex and multifaceted nature of BPD, characterized by significant mood disturbances accompanied by co-manifestation of ADHD.

### White Matter Tract Alterations and association between DTI Metrics and Psychological Scores in BPD

DTI metrics in our study revealed significant differences in white matter tract integrity between BPD patients and HCs. Notably, FA was reduced in several tracts among BPD patients, including the cingulum of the olfactory tract, various regions of the corpus callosum, the fornix, the inferior fronto-occipital fasciculus, and the superior longitudinal fasciculus. These reductions in FA suggest disrupted microstructural integrity and impaired white matter connectivity in these regions. Additionally, increased MD, RD, and AD were observed in the fornix, cingulum of the olfactory tract, corpus callosum (forceps minor), and inferior fronto-occipital fasciculus indicating a potential abnormalities in myelin or axonal properties, further supporting the notion of compromised white matter integrity in BPD. Collectively, our findings underscore the potential role of white matter tract disruptions in the pathophysiology of BPD, potentially contributing to the cognitive and emotional dysregulation observed in affected cases.

While DTI has previously been reruited to study schizophrenic patients, revealing reduced FA in the corpus callosum compared to HCs (3-5), Yurgelun-Todd et al.(6) conducted the first DTI-based study to identify significant differences in corpus callosum microstructure between bipolar patients and age-matched healthy controls. They found that bipolar patients had significantly elevated FA in the genu, but not the splenium, of the corpus callosum compared to the HCs. These results align with earlier reports of white matter macrostructure abnormalities in bipolar disorder (7-10). Additionally, in HCs, FA was higher in the splenium relative to the genu, whereas bipolar patients showed similar FA levels across both regions. Conversely, trace values in the genu were higher than those in the splenium for both patients and controls. This distinct pattern of FA and trace in the anterior (genu) versus posterior (splenium) regions of the corpus callosum is in line with prior studies in HCs (11-13).

Thiel et al (14) recently revealed achieved a differences in white matter microstructure between BPD type I and II, and HCs. Specifically, FA values, were found to be significantly lower in BPD-I patients compared to lack thereof, particularly in the forceps minor of the corpus callosum and other major fiber tracts. This decrease was more pronounced in BD-I than BD-II and HC. Additionally, BD-II patients exhibited lower FA values than HC, albeit to a lesser extent and in smaller clusters within the left body of the CC. The study also found increased RD and MD in BPD-I patients relative to both BPD-II and HC, with no alterations in AD. By contrast, no significant main effect of diagnosis on gray matter volume was achieved. Exploratory analyses suggested potential gray matter volume reductions in BPD-I and-II patients in the parietal, frontal, and parahippocampal/fusiform regions, following the pattern of BD-I < BD-II < HC, though these findings were uncorrected for multiple comparisons. Moreover, the white matter differences between BPD-I and -II remained significant even after adjusting for various clinical variables and polygenic risk scores, indicating robust structural disparities.

Our findings regarding the association between DTI metrics (FA, MD, RD, AD) and psychological assessment scores yielded weak correlation, none of which reached statistical significance. FA and MD exhibited weak positive correlations with neuropsychiatric scales, suggesting that higher white matter integrity or increased diffusion might be associated with more severe symptoms. Conversely, RD and AD showed weak negative correlations, implying that increased diffusion might correlate with milder symptoms. However, the lack of statistical significance indicates that these correlations may not be meaningful or reliable. Consequently, our findings suggest that the observed association between white matter integrity and psychological symptoms should be interpreted with caution, as they may not reflect true associations within this study population.

A decrease in FA, along with an increase in MD, RD, and VR observed in both schizophrenia and BPD by Squarcina et al.(15), may result from disrupted brain connections and/or demyelination. Increases in AD, VR, and MD in BD could suggest tissue structure damage, affecting the boundaries necessary for diffusion. These results are consistent with Benedetti et al. (16), who also reported decreased FA and increased MD and AD in a separate group of BD patients. Additionally, a decrease in FA coupled with an increase in RD, found in the corpus callosum, thalamic radiation, and corona radiata in schizophrenia, as well as in widespread white matter areas in BD, may indicate significant damage to myelin sheaths and barriers perpendicular to the main axis of the axons (17).

Insights from a DTI-based mega- and meta-analyses across 3033 cases by Favre et al(18) investigating white matter microstructural abnormalities in BPD by comparing FA values between 1482 patients with BD and 1551 HC. The mega-analysis revealed that patients with BD had significantly lower FA in 29 out of 43 white matter tracts, particularly in the corpus callosum and bilateral cinguli, indicating widespread microstructural abnormalities. These findings persisted even after accounting for whole-brain average FA, suggesting both global and region-specific white matter disruptions. Age-by-diagnosis interactions showed steeper age-related FA decline in HC for most regions, whereas a different pattern was demonstrated in the cingulum. No robust sex-by-diagnosis interactions were detected. Within the BPD group, earlier age at onset and longer illness duration were associated with lower FA in specific regions. Medication effects were also notable; antipsychotic and anticonvulsant use was linked to lower FA, whereas lithium use was associated with higher FA in several regions. The meta-analysis corroborated these results, showing lower FA in 23 out of 44 regions, with the corpus callosum and cingulum again exhibiting the largest effect sizes. These findings underscore the importance of white matter integrity in BPDs, highlighting specific tracts that may contribute to the disorder’s pathophysiology and could be targets for therapeutic interventions.

### Associations between White Matter Tracts and Brief Psychiatric Rating Scale (BPRS) scores

Our study also explored the associations between DTI metrics and BPRS scores, assessing various symptom dimensions including mania, negative symptoms, positive symptoms, and depression/anxiety. Similar to the previous analyses, the correlations between DTI metrics and BPRS scores were weak and not statistically significant. This suggests that the white matter tract alterations in patients suffering from BPD may not directly correlate with specific symptom dimensions as measured by the BPRS. These findings indicate that while white matter disruptions are evident in BPD, their association to specific clinical symptoms remains poorly understood and warrants further investigation.

A recent study by Squarcina, et al investigating the association between white matter tracts and BPRS scores in BPD, schizophrenia, and HCs. Thirty-three BD patients (18 with type I and 15 with type II), 19 SCZ patients, and 35 HC were recruited. Significant white matter alterations were found in both BD and SCZ patients compared to HCs, with BD patients exhibiting decreased FA and MO in specific regions and increased MD, VR, AD, and RD across widespread areas, including the corpus callosum and internal and external capsules. The study also noted a significant correlation between the duration of illness and FA and VR in BPD patients, suggesting that longer illness duration is associated with greater white matter alterations. However, no direct correlation between BPRS scores and white matter changes was found in BPD patients. These findings highlight the extensive white matter disruption in BPD and its potential association with the chronicity of the disorder, but not necessarily with the severity of current psychiatric symptoms as measured by BPRS.

### Limitations

The authors acknowledge several shortcomings through the study. Firstly, the relatively small sample size, particularly within the BPD group, may reduce the statistical impat of the findings and limit the generalizability of our outcomes. Additionally, the cross-sectional nature of the study precludes any conclusions about causality; longitudinal studies would be necessary to determine whether the observed white matter alterations are a cause or effect of bipolar disorder. The exclusion of potential confounding variables such as medication status is another limitation, as many BPDs may be on psychotropic medications that could affect white matter integrity. Furthermore, the study relies on data from a specific dataset (UCLA CNP), which may have inherent biases or limitations in demographic representation. The lack of significant findings in the correlations between DTI metrics and psychological symptoms may suggests that the relationship between white matter integrity and clinical symptoms may be more complex than captured in the current study, warranting for further inestigations with more comprehensive neuroimaging and clinical assessments.

## Conclusion

The study highlights significant white matter tract alterations in individuals with bipolar disorder compared to healthy controls, suggesting compromised microstructural integrity and connectivity. However, the weak and non-significant correlations between such DTI metrics and various psychological and psychiatric scales indicate that the association between white matter integrity and symptom severity was not fully understood. These findings underscore the need for further research to elucidate the mechanisms underlying white matter disruptions in BPD and their potential impact on clinical outcomes.

## Data Availability

The manuscript provides all the data including analysis and findings.

## References

Altshuler, L. L., Curran, J. G., Hauser, P., Mintz, J., Denicoff, K., & Post, R. (1995). T2 hyperintensities in bipolar disorder: magnetic resonance imaging comparison and literature meta-analysis. The American journal of psychiatry, 152(8), 1139–1144.

Ashburner J. A fast diffeomorphic image registration algorithm. Neuroimage. 2007;38(1):95–113.

Barnea-Goraly, N., Chang, K. D., Karchemskiy, A., & Reiss, A. L. (2009). Limbic and corpus callosum aberrations in adolescents with bipolar disorder: A tract-based spatial statistics analysis. Biological Psychiatry, 66(3), 238–244. 10.1016/j.biopsych.2009.02.027

Beaulieu, C. (2002). The basis of anisotropic water diffusion in the nervous system–a technical review. NMR in Biomedicine: An International Journal Devoted to the Development and Application of Magnetic Resonance In Vivo, 15(7-8), 435–455.

Benedetti, F., Yeh, P. H., Bellani, M., Radaelli, D., Nicoletti, M. A., Poletti, S., … & Smeraldi, E. (2011). Disruption of white matter integrity in bipolar depression as a possible structural marker of illness. Biological Psychiatry, 69(4), 309–317. 10.1016/j.biopsych.2010.07.028

Beyer, J. L., Kuchibhatla, M., Payne, M. E., Moo-Young, M., Cassidy, F., MacFall, J. R., … & Krishnan, K. R. R. (2005). Hippocampal volume measurement in older adults with bipolar disorder. American Journal of Geriatric Psychiatry, 13(5), 407–410. 10.1097/00019442-200505000-00005

Brambilla, P., Nicoletti, M., Sassi, R. B., Mallinger, A. G., Frank, E., Keshavan, M. S., & Soares, J. C. (2004). Corpus callosum signal intensity in patients with bipolar and unipolar disorder. *Journal of Neurology*, Neurosurgery & Psychiatry, 75(2), 221–225.

Buchsbaum, M. S., Tang, C. Y., Peled, S., Gudbjartsson, H., Lu, D., Hazlett, E. A., … & Atlas, S. W. (1998). MRI white matter diffusion anisotropy and PET metabolic rate in schizophrenia. Neuroreport, 9(3), 425–430.

Cao, X., Yang, Y., Zhou, X., et al. (2024). White matter abnormalities associated with depressive and psychotic symptoms in bipolar disorder. Neuropsychopharmacology. 10.1038/s41386-024-01812-7

Chepuri, N. B., Yen, Y. F., Burdette, J. H., Li, H., Moody, D. M., & Maldjian, J. A. (2002). Diffusion anisotropy in the corpus callosum. American journal of Neuroradiology, 23(5), 803–808.

Dahnke, R., Yotter, R. A., & Gaser, C. (2013). Cortical thickness and central surface estimation. Neuroimage, 65, 336–348.

Duarte, J. A., Massuda, R., Goi, P. D., Vianna-Sulzbach, M., Colombo, R., Kapczinski, F., & Gama, C. S. (2018). White matter volume is decreased in bipolar disorder at early and late stages. Trends in Psychiatry and Psychotherapy, 40(4), 277–284. 10.1590/2237-6089-2017-0025

El Nagar, Z. M., El Shahawi, H. H., Effat, S. M., El Sheikh, M. M., Adel, A., Ibrahim, Y. A., & Aufa, O. M. (2022). Single episode brief psychotic disorder versus bipolar disorder: A diffusion tensor imaging and executive functions study. Schizophrenia Research: Cognition, 27, 100214.

Ellison-Wright, I., & Bullmore, E. (2010). Anatomy of bipolar disorder and schizophrenia: a meta-analysis. Schizophrenia research, 117(1), 1–12.

Ertek, I. E., Ilhan, M. N., Dikmen, A. U., & Gozukara, M. (2020). Is There A Relationship Between Adult ADHD and Bipolar Symptoms? A Cross-Sectional Study with Primary Care Applicants in Ankara, Turkey. Psychiatry and Clinical Psychopharmacology, 30(4), 374–380.

Favre, P., Pauling, M., Stout, J., Hozer, F., Sarrazin, S., Abé, C., … & ENIGMA Bipolar Disorder Working Group. (2019). Widespread white matter microstructural abnormalities in bipolar disorder: evidence from mega-and meta-analyses across 3033 individuals. Neuropsychopharmacology, 44(13), 2285–2293.

Foong, J., Maier, M., Clark, C. A., Barker, G. J., Miller, D. H., & Ron, M. A. (2000). Neuropathological abnormalities of the corpus callosum in schizophrenia: a diffusion tensor imaging study. *Journal of Neurology*, Neurosurgery & Psychiatry, 68(2), 242–244.

GBD 2019 Mental Disorders Collaborators. (2022). Global, regional, and national burden of 12 mental disorders in 204 countries and territories, 1990–2019: a systematic analysis for the Global Burden of Disease Study 2019. The Lancet Psychiatry.

Gorgolewski, K. J., Durnez, J., & Poldrack, R. A. (2017). Preprocessed consortium for neuropsychiatric phenomics dataset. F1000Research, 6, 1262.

Haghshomar, M., Rahmani, F., Aarabi, M. H., Shahjouei, S., Sobhani, S., & Rahmani, M. (2019). White matter changes correlates of peripheral neuroinflammation in patients with Parkinson’s disease. Neuroscience, 403, 70–78.

Hamilton, M. (1960). A rating scale for depression. Journal of neurology, neurosurgery, and psychiatry, 23(1), 56.

Haznedar, M. M., et al. (2005). Frontal white matter and cingulum abnormalities in schizophrenia: A diffusion tensor imaging study. Biological Psychiatry, 57(7), 727–734. 10.1016/j.biopsych.2004.12.008

Jiang, Y., Luo, C., Li, X., Li, Y., Yang, H., Li, J., … & Yao, D. (2019). White-matter functional networks changes in patients with schizophrenia. Neuroimage, 190, 172–181.

Jonathan, G. K., Dopke, C. A., Michaels, T., Bank, A., Martin, C. R., Adhikari, K., … & Goulding, E. H. (2021). A smartphone-based self-management intervention for bipolar disorder (LiveWell): User-centered development approach. JMIR Mental Health, 8(4), e20424. 10.2196/20424

Lee, D. K., Lee, H., Ryu, V., Kim, S. W., & Ryu, S. (2022). Different patterns of white matter microstructural alterations between psychotic and non-psychotic bipolar disorder. PLoS ONE, 17(3 March), 1–13. 10.1371/journal.pone.0265671

Li, T., Li, R., Zhao, L., Sun, Y., Wang, C., & Bo, Q. (2024). Comparative Analysis of Personality Traits in Major Depressive Disorder and Bipolar Disorder: Impact, Differences, and Associations with Symptoms. Neuropsychiatric Disease and Treatment, 363–371.

Lim, K. O., Hedehus, M., Moseley, M., De Crespigny, A., Sullivan, E. V., & Pfefferbaum, A. (1999). Compromised white matter tract integrity in schizophrenia inferred from diffusion tensor imaging. Archives of general psychiatry, 56(4), 367–374.

Linke, J. O., Stavish, C., Adleman, N. E., Sarlls, J., Towbin, K. E., Leibenluft, E., & Brotman, M. A. (2020). White matter microstructure in youth with and at risk for bipolar disorder. Bipolar disorders, 22(2), 163–173.

Liu, H., Yang, Y., Xia, Y., Zhu, W., Leak, R. K., Wei, Z., … & Hu, X. (2017). Aging of cerebral white matter. Ageing Research Reviews, 34, 64–76. 10.1016/j.arr.2016.11.005

Magioncalda, P., Martino, M., Conio, B., Piaggio, N., Teodorescu, R., Escelsior, A., … & Inglese, M. (2016). Patterns of microstructural white matter abnormalities and their impact on cognitive dysfunction in the various phases of type I bipolar disorder. Journal of Affective Disorders, 193, 39–50. 10.1016/j.jad.2015.12.066

Maller, J. J., Thaveenthiran, P., Thomson, R. H., McQueen, S., & Fitzgerald, P. B. (2014). Volumetric, cortical thickness and white matter integrity alterations in bipolar disorder type I and II. Journal of affective disorders, 169, 118–127.

Marlinge, E., Bellivier, F., & Houenou, J. (2014). White matter alterations in bipolar disorder: potential for drug discovery and development. Bipolar Disorders, 16(2), 97–112. 10.1111/BDI.12135.

McGuffin, P., Rijsdijk, F., Andrew, M., Sham, P., Katz, R., & Cardno, A. (2003). The heritability of bipolar affective disorder and the genetic relationship to unipolar depression. Archives of General Psychiatry, 60(5), 497–502. 10.1001/archpsyc.60.5.497

Merikangas, K. R., Akiskal, H. S., Angst, J., Greenberg, P. E., Hirschfeld, R. M., Petukhova, M., & Kessler, R. C. (2007). Lifetime and 12-month prevalence of bipolar spectrum disorder in the National Comorbidity Survey replication. Archives of General Psychiatry, 64(5), 543–552. 10.1001/archpsyc.64.5.543

Murphy, C. E., Walker, A. K., & Weickert, C. S. (2021). Neuroinflammation in schizophrenia: the role of nuclear factor kappa B. Translational psychiatry, 11(1), 528.

Oertel-Knöchel, V., Reinke, B., Alves, G., Jurcoane, A., Wenzler, S., Prvulovic, D., Linden, D., & Knöchel, C. (2014). Frontal white matter alterations are associated with executive cognitive function in euthymic bipolar patients. Journal of Affective Disorders, 155(1), 223–233. 10.1016/j.jad.2013.11.004.

Patel, R. S., Virani, S., Saeed, H., Nimmagadda, S., Talukdar, J., & Youssef, N. A. (2018). Gender differences and comorbidities in U.S. adults with bipolar disorder. Brain Sciences, 8(9), 168. 10.3390/brainsci8090168

Pfefferbaum, A., & Sullivan, E. V. (2003). Increased brain white matter diffusivity in normal adult aging: relationship to anisotropy and partial voluming. Magnetic Resonance in Medicine: An Official Journal of the International Society for Magnetic Resonance in Medicine, 49(5), 953–961.

Poldrack, R. A., Congdon, E., Triplett, W., Gorgolewski, K. J., Karlsgodt, K. H., Mumford, J. A., … & Bilder, R. M. (2016). A phenome-wide examination of neural and cognitive function. Scientific data, 3(1), 1–12.

Radoeva, P. D., Jenkins, G. A., Schettini, E., Gilbert, A. C., Barthelemy, C. M., DeYoung, L. L. A., Kudinova, A. Y., Kim, K. L., MacPherson, H. A., & Dickstein, D. P. (2020). White matter correlates of cognitive flexibility in youth with bipolar disorder and typically developing children and adolescents. Psychiatry Research - Neuroimaging, 305(February). 10.1016/j.pscychresns.2020.111169

Safaiyan, S., Besson-Girard, S., Kaya, T., Cantuti-Castelvetri, L., Liu, L., Ji, H., … & Simons, M. (2021). White matter aging drives microglial diversity. Neuron, 109(7), 1100– 1117. 10.1016/j.neuron.2021.01.027

Safaiyan, S., Besson-Girard, S., Kaya, T., Cantuti-Castelvetri, L., Liu, L., Ji, H., … & Simons, M. (2021). White matter aging drives microglial diversity. Neuron, 109(7), 1100–1117. DOI: 10.1016/j.neuron.2021.01.027

Santos, J. P. L., Versace, A., Stiffler, R. S., Aslam, H. A., Lockovich, J. C., Bonar, L., Bertocci, M., Iyengar, S., Bebko, G., & Skeba, A. (2022). White matter predictors of worsening of subthreshold hypomania severity in non-bipolar young adults parallel abnormalities in individuals with bipolar disorder. Journal of Affective Disorders, 306, 148–156. DOI: 10.1016/j.jad.2022.03.039

Schneider, J. F., Il’Yasov, K. A., Hennig, J., & Martin, E. (2004). Fast quantitative diffusion-tensor imaging of cerebral white matter from the neonatal period to adolescence. Neuroradiology, 46, 258–266.

Silverstone, T., McPherson, H., Li, Q., & Doyle, T. (2003). Deep white matter hyperintensities in patients with bipolar depression, unipolar depression and age-matched control subjects. Bipolar disorders, 5(1), 53–57.

Smith, S. M., Jenkinson, M., Woolrich, M. W., Beckmann, C. F., Behrens, T. E. J., Johansen-Berg, H., … & Matthews, P. M. (2004). Advances in functional and structural MR image analysis and implementation as FSL. NeuroImage, 23(S1), S208–S219. 10.1016/j.neuroimage.2004.07.051

Squarcina, L., Bellani, M., Rossetti, M. G., Perlini, C., Delvecchio, G., Dusi, N., … & Brambilla, P. (2017). Similar white matter changes in schizophrenia and bipolar disorder: A tract-based spatial statistics study. PLoS One, 12(6), e0178089.

Stephen, R., Solomon, A., Ngandu, T., Levälahti, E., Rinne, J. O., Kemppainen, N., … & FINGER Study Group. (2020). White matter changes on diffusion tensor imaging in the FINGER randomized controlled trial. Journal of Alzheimer’s Disease, 78(1), 75–86.

Strakowski, S. M., Wilson, D. R., Tohen, M., Woods, B. T., Douglass, A. W., & Stoll, A. L. (1993). Structural brain abnormalities in first-episode mania. Biological psychiatry, 33(8-9), 602–609.

Strakowski, S. M., Adler, C. M., Almeida, J., Altshuler, L. L., Blumberg, H. P., Chang, K. D., … & DelBello, M. P. (2012). The functional neuroanatomy of bipolar disorder: a consensus model. Bipolar Disorders, 14(4), 313–325. 10.1111/j.1399-5618.2012.01022.x

Thiel, K., Lemke, H., Winter, A., Flinkenflügel, K., Waltemate, L., Bonnekoh, L., … & Dannlowski, U. (2024). White and gray matter alterations in bipolar I and bipolar II disorder subtypes compared with healthy controls–exploring associations with disease course and polygenic risk. Neuropsychopharmacology, 49(5), 814–823.

Vieta, E., Berk, M., Schulze, T. G., Carvalho, A. F., Suppes, T., Calabrese, J. R., … & Grande, I. (2018). Bipolar disorders. Nature Reviews Disease Primers, 4(1), 18008. 10.1038/nrdp.2018.8

Yotter, R. A., Dahnke, R., Thompson, P. M., & Gaser, C. (2011). Topological correction of brain surface meshes using spherical harmonics. Human brain mapping, 32(7), 1109–1124.

Yurgelun-Todd, D. A., Silveri, M. M., Gruber, S. A., Rohan, M. L., & Pimentel, P. J. (2007). White matter abnormalities observed in bipolar disorder: a diffusion tensor imaging study. Bipolar disorders, 9(5), 504–512.

